# Patient Risk-Benefit Preferences for Transcatheter versus Surgical Mitral Valve Repair

**DOI:** 10.1101/2023.05.24.23290501

**Authors:** Anna Hung, Jui-Chen Yang, Matthew Wallace, Brittany A. Zwischenberger, Sreekanth Vemulapalli, Robert J. Mentz, Elizabeth Thoma, Scott Goates, John Lewis, Susan Strong, Shelby D. Reed

**Affiliations:** Duke Clinical Research Institute, Duke University School of Medicine, Durham NC, United States; Center of Innovation to Accelerate Discovery and Practice Transformation, Durham Veterans Affairs Health Care System, Durham, NC, United States; Department of Population Health Sciences, Duke University School of Medicine, Durham, NC, United States; Division of Cardiovascular and Thoracic Surgery, Department of Surgery, Duke University Medical Center, Durham, NC, United States; Division of Cardiology, Department of Medicine, Duke University Medical Center, Durham, NC, United States; Abbott Laboratories, Chicago, IL, United States; Heart Valve Voice US, Washington, DC, United States

**Keywords:** Mitral regurgitation, transcatheter mitral valve repair procedure, mitral valve repair surgery, transcatheter edge-to-edge repair

## Abstract

**BACKGROUND:** Transcatheter repair of mitral regurgitation (MR) is less invasive than surgery, but has greater five-year mortality and re-intervention risks, as well as more limited functioning. The study objective was to quantify patient preferences for risk-benefit tradeoffs associated with transcatheter and surgical options for MR.

**METHODS:** A discrete-choice experiment survey was administered to patients with MR recruited through a patient advocacy organization. Attributes (and levels) included: procedure type (transcatheter versus surgical); risk of 30-day mortality (2%, 5%, and 10%); risk of five-year mortality (20%, 30%, and 45%) and physical functioning for five years (corresponding to improvements from New York Heart Association [NYHA] class III to I or class III to II); number of hospitalizations (1, 4, and 8) in the next five years; and risk of additional surgery in the next five years (10%, 20%, and 30% or 40%). A mixed-logit regression model was fit to estimate preference weights.

**RESULTS:** 201 individuals completed the survey: 63% were female; mean age was 74 years. On average, respondents preferred transcatheter repair over surgery. To undergo a less invasive procedure (i.e., transcatheter repair), respondents would accept up to a 13.3% (95% confidence interval [CI]: 8.7% to 18.5%) increase in re-intervention risk above a baseline of 10%, 4.6 (95% CI: 3.1 to 6.2) more hospitalizations above a baseline of one, a 9.3% (95% CI: 5.2% to 14.3%) increase in mortality risk above a baseline of 20%, or more limited physical functioning representing nearly one NYHA class (0.8, 95% CI: 0.5 to 1.3) over five years.

**CONCLUSIONS:** Patients in general preferred a transcatheter procedure over surgery. When holding constant all other factors, a functional improvement from NYHA class III to class I maintained over five years would be needed, on average, for patients to prefer surgery over a transcatheter procedure.

## INTRODUCTION

Mitral regurgitation (MR) is the most common valvular heart disease in the United States, affecting approximately 10% of adults 75 years of age and above.^1^ MR is more common in older adults, and as the population ages, the number of mitral valve repair interventions is expected to increase.^1, 2^ For patients with severe degenerative MR, two options for mitral valve repair include surgery and transcatheter edge-to-edge repair (TEER).^3^

Compared to surgery, TEER is less invasive and requires a shorter recovery time. In a randomized clinical trial, TEER patients also had a slightly lower risk of 30-day mortality compared to surgery patients (1.1% versus 2.1%, respectively).^4^ However, compared to surgery patients, TEER patients also had a higher incidence of re-intervention over the next five years (27.9% versus 8.9%), a higher five-year mortality risk (26.8% versus 20.8%), and more limited physical functioning (8.6% versus 2.5% had New York Heart Association (NYHA) class III or IV symptoms at the end of five years, and approximately 65% versus nearly 80% had NYHA class I symptoms at the end of five years).^5^ These findings were based on a single trial that enrolled patients from 2005 to 2008. Whether these findings remain true with contemporary standards and practices is unknown. To fill this gap, two clinical trials comparing transcatheter versus surgical interventions for patients with severe degenerative MR are currently underway.^6, 7^ When results become available, it will be important to understand patient acceptability of tradeoffs between benefits and risks of the two intervention types to help inform treatment decisions.

The objective of this study was to quantify patients’ stated preferences related to risk-benefit tradeoffs associated with a transcatheter procedure, like TEER, versus surgery. Specifically, the goal was to quantify the maximum-acceptable risk (MAR) patients were willing to accept to undergo a less invasive procedure (i.e., a transcatheter procedure) rather than surgery for MR. A secondary objective was to explore heterogeneity in patient preferences.

## METHODS

A discrete-choice experiment (DCE) survey was designed according to best practices and guidance issued by the United States Food and Drug Administration Center for Devices and Radiological Health and stated-preference experts.^8–11^ The DCE approach was selected among several stated preference approaches because of its rigor when quantifying patient risk-benefit tradeoffs through a series of choice questions and its flexibility in evaluating a range of potentially relevant tradeoffs.^8^

### DCE Survey Design

Each of the choice questions (example shown in Figure 1) in this survey featured a choice between two hypothetical MR interventions. Survey participants were asked to choose a procedure, assuming this was their initial procedure and assuming they had daily physical function limitations similar to NYHA class III. Survey participants were shown 12 choice questions, each showing different profiles for the two interventions in terms of key features (i.e., attributes). The attributes and levels (i.e., possible values shown for each attribute) are listed in Table 1 and included: intervention type (transcatheter versus surgical); risk of 30-day mortality (2%, 5%, and 10%); risk of five-year mortality (20%, 30%, and 45%) and level of physical functioning for those five years (corresponding to improvements from NYHA class III to class I or from class III to class II); number of hospitalizations (1, 4, and 8) in the next five years; and risk of additional surgery in the next five years (10%, 20%, and 30% or 40%). To emphasize the interdependency between the duration of improvements in physical functioning (up to five years) and five-year mortality risk, these two attributes were presented as a compound attribute. The attributes and levels were informed by a literature review of common clinical trial endpoints for TEER versus surgery,^4, 5, 12–14^ discussion with a patient advisory group of six patients with MR (or history of MR), and discussion with the study team that included practicing cardiologists and a cardiothoracic surgeon.

**Figure 1.**
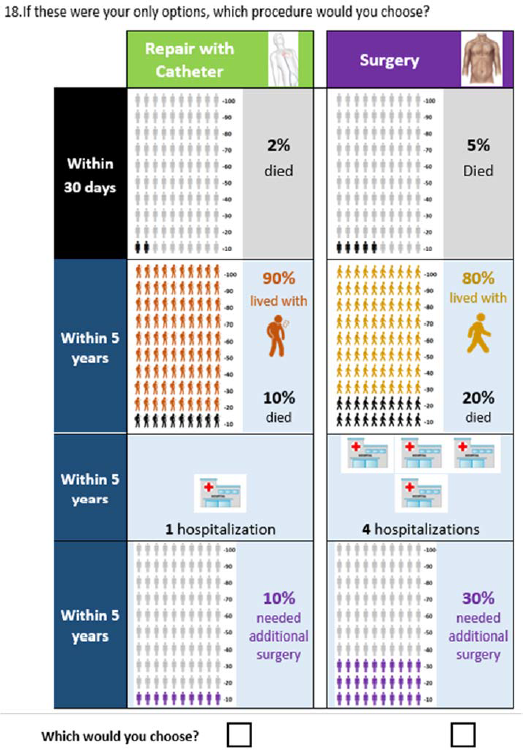
Example Discrete Choice Question.

**Table 1.**
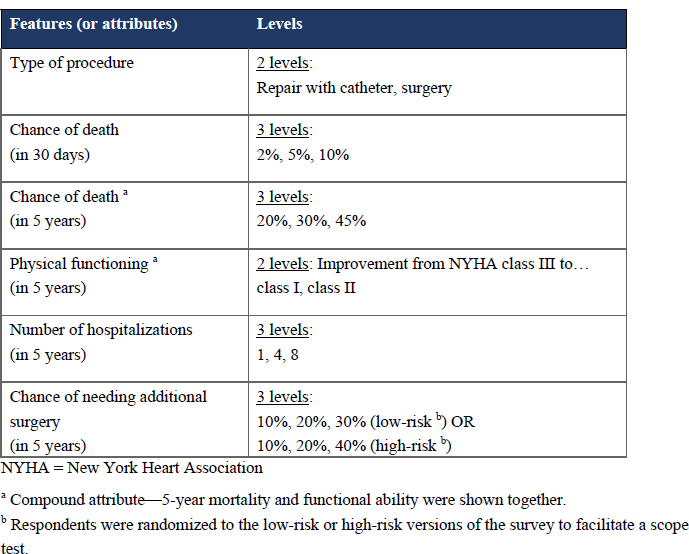
Attributes and Levels.

Prior to answering these choice questions, the survey provided educational material, including descriptive information about each of the attributes and graphical depictions (e.g., icon arrays to help survey participants understand probabilities). The survey was pretested with nine individuals with MR recruited through clinician collaborators and Heart Valve Voice US, a patient-led non-profit organization that advocates for patients with heart valve disease. During one-on-one pretest interviews, the survey instrument was assessed for: comprehensibility and readability, that sufficient information was provided for each of the attributes, and that the ranges of attribute levels were sufficiently broad to encourage participants to make tradeoffs among attributes. Iterative improvements to the survey instrument were made based on these interviews. The survey also assessed the participants’ understanding of the content using multiple-choice comprehension questions and participants’ choice consistency and logic using several internal validity tests. One of the internal validity tests was a scope test in which respondents were randomized to two different versions of the survey. In one version, the highest level shown for the chance of needing additional surgery was 30% (therefore, the set of possible levels was 10%, 20%, and 30%) and in the other version, the highest level shown was 40% (therefore, the set of possible levels was 10%, 20%, and 40%). In the analysis stage, this allowed for an assessment of whether participants accounted for the numerical value rather than mentally re-recoding the levels as ‘low’, ‘medium’, and ‘high’ in which case a difference in preference weights between 30% and 40% levels would not be expected.

Lastly, the survey collected demographic and medical information from participants. Since the survey included detailed information regarding how the MR interventions would be performed and multiple attributes related to risk, participants’ health literacy and numeracy were measured in the survey using the validated Brief Health Literacy Screen (BHLS)^15, 16^ and the validated, three-item version of the Subjective Numeracy Scale (SNS-3)^17, 18^.

### Experimental Design

An experimental design was created using SAS statistical software version 9.4 (SAS Institute, Cary, NC) to determine the combinations of attribute levels shown for each hypothetical procedure profile and all pairs of hypothetical procedure profiles that populated the choice questions. Three hundred and sixty choice questions were constructed according to a D-efficient experimental design per best practices and divided into 45 blocks of eight choice questions and 90 blocks of four choice questions.^9^ Each survey participant was randomly assigned to a set of 12 choice questions from the two blocks (representing the first eight and then final four choice questions). This design provided a safeguard in case survey participants did not answer all 12 choice questions. Additionally, one-quarter of the choice questions displayed the same intervention type for both profiles and one-quarter of the choice questions displayed the same 5-year risk of mortality for both profiles to ensure that additional preference information could be obtained among participants whose choices were driven by their preferred intervention type or lowest 5-year mortality risk.

### Data Collection

The survey was programmed in Lighthouse Studio version 9.12.0 (Sawtooth Software, Salt Lake City, UT) for web-based administration. Eligible participants included adults 18 years of age and older who reported a physician diagnosis of MR. Participants were recruited from November 1, 2021 to July 4, 2022 by Heart Valve Voice US, which targeted their patient community and arranged social media campaigns. During recruitment, it was identified that women predominated the study sample. To address this imbalance, starting on May 3, 2022, recruitment was restricted to men. Prior to accessing the survey, participants were required to give informed consent. The study was approved by the institutional review board for Duke University Health System.

### Statistical analysis

The response data were assessed for data quality and analyzed according to an *a priori* statistical analysis plan. Prior to analysis, the data quality of response data was assessed through a series of internal validity tests,^19^ such as checking to ensure survey respondents did not always choose the option on the left or right (i.e., straightlining) and assessing whether survey respondents always made a choice based on a single attribute (i.e., attribute dominance). Another internal validity test consisted of a choice question in which one of the two options was better across all attributes, and therefore served as a test that respondents understood the choice question layout (Supplemental Appendix Figure 1). Additional checks included assessing time to complete the survey (i.e., if surveys were completed very quickly, such as less than five minutes) and performance on comprehension questions (i.e., how many of the eight comprehension questions were answered incorrectly). Evaluation of data quality was conducted using MATLAB version R2022a (The Mathworks, Inc., Natick, MA) and SAS statistical software version 9.4 (SAS Institute, Cary, NC).

After excluding any survey responses that were not deemed to be internally valid, performance on the scope test was assessed (details provided in the Supplemental Appendix) to determine whether survey response data could be combined across the two versions. Random-parameters logit models were fit to choice data to calculate preference weight estimates for each of the attribute levels, and tests for interactions between the two attributes in the compound attribute (five-year mortality risk and physical functioning) were performed. Next, the mean maximum acceptable increase in five-year risk of needing additional surgery that respondents would be willing to accept for a less invasive procedure (i.e., transcatheter procedure instead of surgery) was calculated by determining the increase in preference utility for a less invasive procedure and then determining the amount of additional risk that would lead to an equivalent decrease in preference utility. Similarly, the mean maximum acceptable increases in number of hospitalizations, mortality risk, and limited physical functioning for a transcatheter procedure instead of surgery were calculated, and 95% confidence intervals were generated using the Krinsky-Robb procedure.^20^ Analyses were conducted in Stata SE version 16 (StataCorp LP, College Station, TX). In addition, simultaneous MARs for five-year mortality and needing additional surgery were calculated using Microsoft Excel (Microsoft, Redmond, WA).^21^

Lastly, to evaluate heterogeneity in patient preferences, latent-class analyses were performed to identify preference subgroups. The final model (and number of latent classes) was identified based on evaluation of data-driven criteria including Bayesian information criterion, Akaike information criterion, and entropy as well as subjective criteria ensuring that preferences were different between latent classes (i.e., that there were not so many latent classes identified that preferences were not different). Covariates included in these latent class analyses were determined *a priori* and included: demographics such as older age (≥80), gender, and marital status; lower literacy (BHLS <10) and lower numeracy (SNS <13), with cutpoints determined based on lowest quartile; clinical characteristics such as more severe disease (defined as reporting more than two of the past seven days with NYHA Class III symptoms, with the cutpoint determined based on the highest quartile), prior surgery, prior transcatheter procedure, prior complication (after surgery or transcatheter procedure), and concomitant heart failure; and performance on internal validity checks such as answering the dominated-pair choice question incorrectly, answering at least half of the comprehension questions incorrectly, and a shorter survey completion time (<15 minutes, representing less than 5% of the sample). Analyses were conducted in LatentGOLD version 6.0 (Statistical Innovations Inc, Arlington, MA).

## RESULTS

### Patient Demographic and Medical Characteristics

Of the 201 survey respondents, the average age was 73.7 years and 62.8% were women (Table 2). The majority were white (94.6%) and over half were married (64.5%). The sample represented mostly retired individuals (80.1%; Supplemental Table 1) and 49.4% had at least a four-year college degree. Subjective health literacy and numeracy were high (mean BHLS score of 10.4 with a maximum health literacy score of 15 and mean SNS-3 score of 15.0 with a maximum numeracy score of 18). Common comorbidities included hypertension (54.7%), atrial fibrillation (45.8%), and heart failure (30.8%). Nearly a quarter (24.4%) reported having a prior mitral valve surgery and 13.9% reported having a prior mitral valve transcatheter procedure.

**Table 2.**
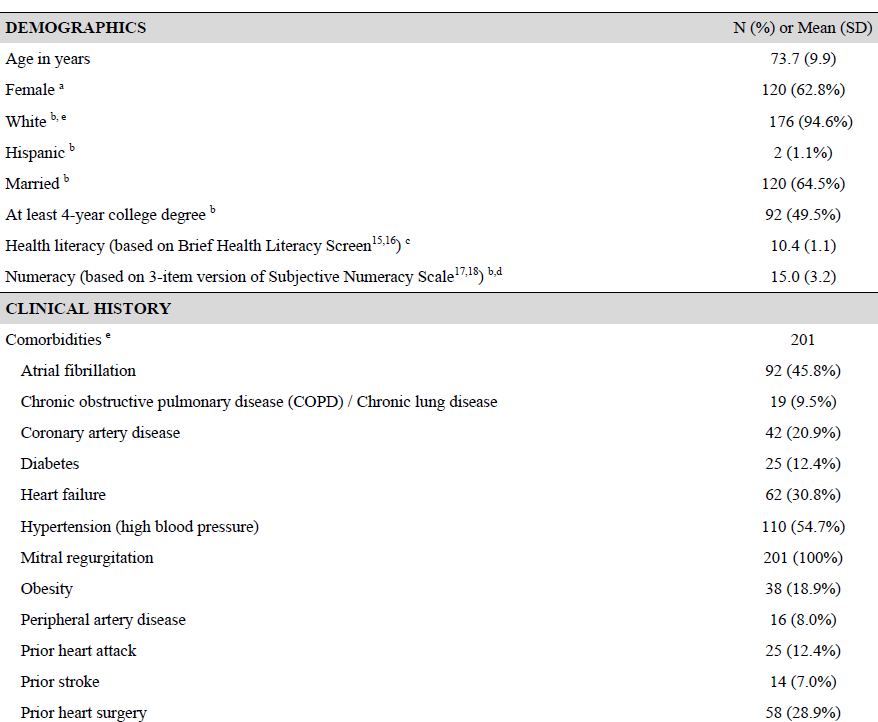
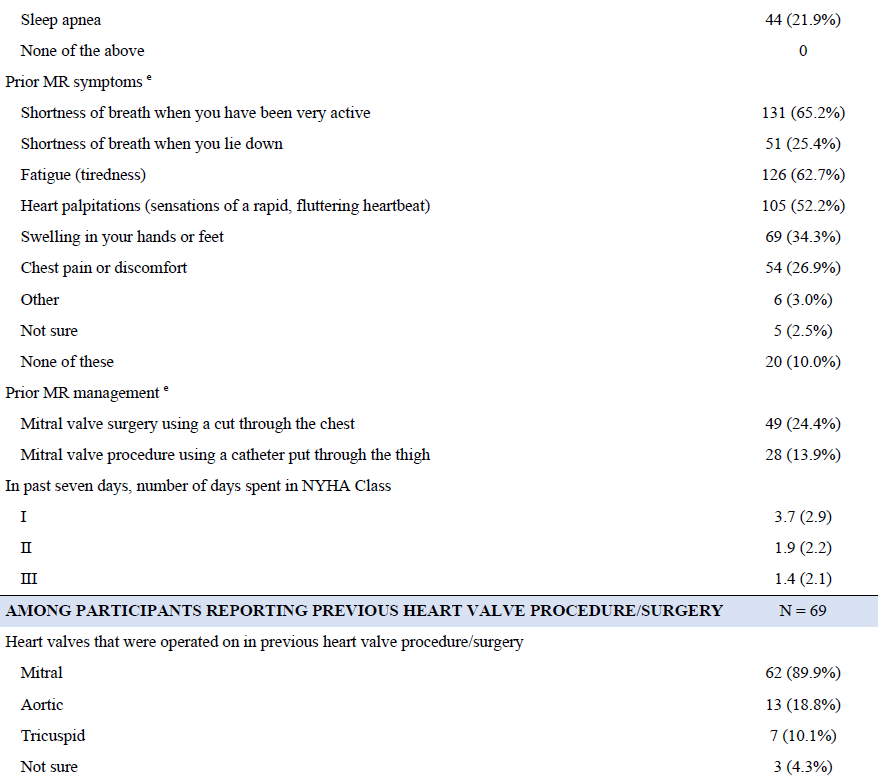
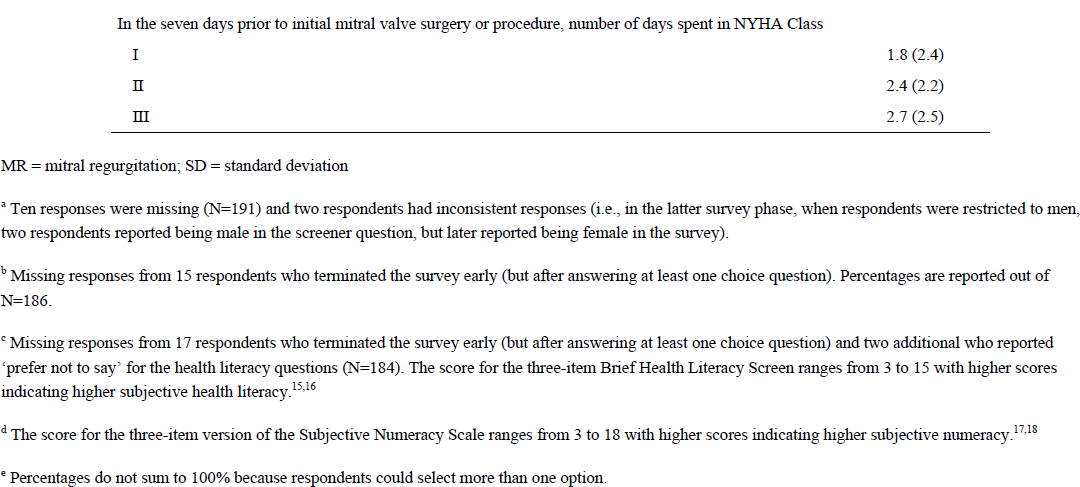
Respondent Characteristics (N=201)

### Data Quality

Performance on internal validity tests generally indicated that the respondents were attentive and understood the choice tasks. No respondents finished the survey in less than five minutes, and no respondents answered all of the eight comprehension questions incorrectly. Less than 10% dominated on a single attribute, and 7% answered the dominated-pair choice question incorrectly. Based on the scope test, participants were paying attention to the numeric value (30% versus 40%, depending on randomized survey version) of the highest level considered for chance of needing additional surgery evidenced by more negative utility weights estimated for 40% as compared to 30% risk levels, allowing for the pooling of response data for analysis. The Supplemental Appendix provides full details on assessment of data quality.

One exception to the excellent performance on internal validity tests was two respondents who always chose the procedure option on the left or always chose the procedure option on the right across all twelve choice questions (i.e., straightlined). Since the chance of these participants paying attention to the survey was very low, their response data were excluded from the final analytic sample (n=199).

### Preference weight estimates

On average, respondents significantly preferred a transcatheter procedure over surgery as indicated by a positive preference weight for “catheter” and a negative preference weight for “surgery” (p<0.001) (Figure 2, Supplemental Tables 2a-2b). Preferences for all risks and number of hospitalizations were logically ordered, with larger preference weights for lower risks and fewer hospitalizations. Preference weights were also significantly larger for MR interventions that could improve patients’ physical functioning from a level equivalent to NYHA class III to NYHA class I (green) relative to improvements from NYHA class III to class II (orange) across varying five-year mortality risks.

**Figure 2.**
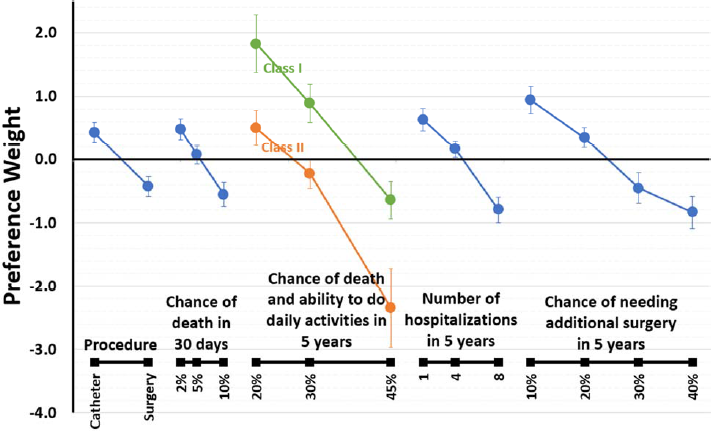
Preference Weight Estimates from Overall Sample (N=199)

### Benefit-risk Tradeoffs

To undergo a less invasive procedure (i.e., a transcatheter procedure), respondents would accept over the next five years up to either (Table 3):

- a 13.3% (95% confidence interval [CI]: 8.7% to 18.5%) increase in risk of re-intervention surgery above a baseline risk of 10%,
- 4.6 (95% CI: 3.1 to 6.2) more hospitalizations above a baseline of one hospitalization,
- a 9.3% (95% CI: 5.2% to 14.3%) increase in mortality risk above a baseline risk of 20%, assuming functional improvement from NYHA class III to class I over those five years, or
- more limited physical functioning representing an equivalent of nearly one NYHA class (0.8, 95% CI: 0.5 to 1.3), assuming a five-year mortality risk of 30%.
- Conversely, respondents’ choices indicated that they would undergo surgery if five-year gains in physical functioning were equivalent to achieving NYHA class I from a baseline of NYHA class III (as opposed to achieving NYHA class II).

**Table 3.** Maximum Acceptable Harm Estimates for a Less Invasive Procedure (Transcatheter Procedure instead of Surgery)

| Harm | Mean Estimate (95% Confidence Interval) |  |
| --- | --- | --- |
| Risk of needing additional surgery (above a baseline risk of 10%) | 13.3% (8.7% - 18.5%) |  |
| Number of hospitalizations (above a baseline risk of one hospitalization) | 4.6 (3.1 - 6.2) |  |
| Risk of 5-year mortality (above a baseline risk of 20%) | 9.3% (5.2% - 14.3%) with improvement from NYHA class III to I | 10.7% (6.5% - 14.5%) with improvement from NYHA class III to II |
| More limited physical functioning (of the difference between NYHA class I and II) | 0.8 (0.5 to 1.3) with 30% five-year mortality risk | 0.7 (0.4 to 1.1) with 20% five-year mortality risk |

NYHA = New York Heart Association

For the compound attribute, when assuming a smaller functional improvement (an improvement of NYHA class III to II), the maximum-acceptable increase in five-year risk of death did not significantly change (10.7%, 95% CI: 6.5% to 14.5%). Similarly, when assuming a smaller five-year risk of death (20%), respondents were willing to accept slightly smaller improvements in physical functioning (0.7, 95% CI: 0.4 to 1.1).

Figure 3 shows the MAR thresholds when considering two risks, five-year mortality (assuming improvement from a baseline of NYHA class III to class II) and re-intervention surgery, simultaneously. When considering these two risks simultaneously, respondents would be willing to accept various combinations of five-year mortality and re-intervention surgery risk (e.g., a 8%-point increase above a baseline of 20% (or 28%) and a 5%-point increase above a baseline of 10% (or 15%), respectively; or 24% and 20%, respectively; and so forth) for a less invasive procedure (transcatheter repair versus surgery).

**Figure 3.**
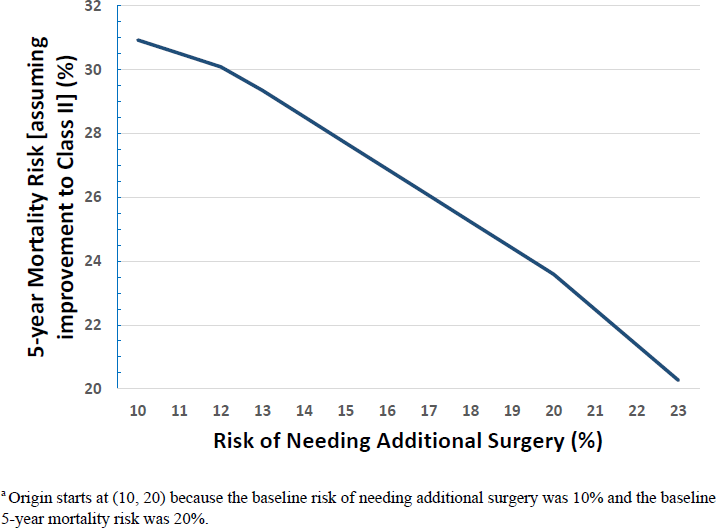
Simultaneous Maximum-Acceptable Risk Thresholds for 5-year Mortality and Needing Additional Surgery for a Less Invasive Procedure (Transcatheter Repair versus Surgery)^a^.

### Preference subgroups

Based on latent-class analyses, four preference subgroups were identified (Figure 4): (1) latent class 1 (representing approximately 33% of the sample), who cared most about functional improvement; (2) latent class 2 (representing approximately 31% of the sample), who cared most about five-year risk of mortality and secondarily, chance of needing additional surgery; (3) latent class 3 (representing approximately 28% of the sample), who cared most about receiving a transcatheter procedure (over surgery); and (4) latent class 4 (representing approximately 8% of the sample), who had disordered preferences and when compared to Latent Class 1, failed internal validity tests, had lower literacy, and answered at least 50% of the comprehension questions incorrectly (Table 4; p-values all <0.05). Compared to Latent Class 1, members of Latent Class 2 had lower literacy and members of Latent Class 3 had lower numeracy, more severe disease, and no prior surgery.

**Figure 4.**
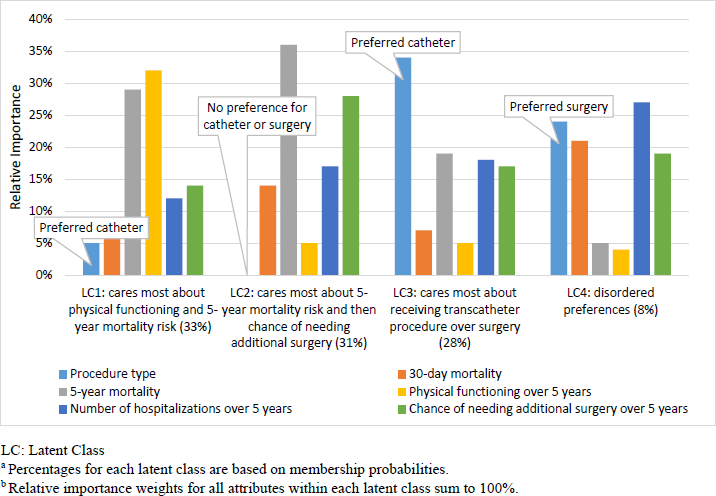
Relative Importance of Attributes by Latent Class (Overall Sample, N=199) ^a,b^.

**Table 4.** Respondent Characteristics Associated with Latent Class Membership as Compared to Latent Class 1 (Overall sample, N=199)

| **Latent Class 2 (Most Concerned with 5-year Mortality Risk)** |  |
| --- | --- |
| **Negative** | **Positive** |
|  | Lower literacy* |
| **Latent Class 3 (Greatest Preference for Transcatheter Procedure)** |  |
| **Negative** | **Positive** |
| Prior surgery* | Lower numeracy** |
|  | More severe disease** |
| **Latent Class 4 (Disordered Preferences)** |  |
| **Negative** | **Positive** |
| More severe disease* | Failed dominating test** |
|  | Got 50%+ of quiz questions wrong** |
|  | Lower literacy** |
|  | Prior surgery* |

\*p<0.1; \*\*p<0.05
Lower literacy and numeracy were defined based on lower scores (less than quartile one) for the Brief Health Literacy Screen<sup>15,16</sup> and three-item Subjective Numeracy Scale<sup>17,18</sup>.
More severe disease was defined based on reporting at least 3 of the 7 days with NYHA Class III level of functioning.
Additional characteristics included that were not significant predictors of class membership:
Age ≥80, female, married, prior transcatheter repair, prior complication (after surgery or transcatheter repair), heart failure, and survey time < 15 min

## DISCUSSION

This DCE study found that patients with MR in general have a preference for a transcatheter procedure over surgery and quantified the amount of potential longer-term risks that patients would accept to opt for a transcatheter procedure. All else equal, in the five years after the intervention, patients were willing to accept any one of the following tradeoffs: an average absolute increase of over 10% in risk of re-intervention (above a baseline risk of 10%); an average increase of about five hospitalizations (above a baseline risk of one hospitalization); an average absolute increase of almost 10% in risk of death (above a baseline risk of 20%); or more limited physical functioning equivalent of improvement from NYHA class III to NYHA class II instead of an improvement to class I. This also means that holding all other factors constant, patients would be willing to undergo surgery if surgery allowed them to improve in physical functioning from the equivalent of NYHA class III to class I, and this improvement were maintained over five years.

How these patients’ risk-benefit acceptability thresholds compare to updated evidence between TEER and surgery remains to be seen. The two global clinical trials that are underway started in 2020 and 2022, respectively, and are estimated to have primary outcome data collected by February 2024 and January 2028, respectively.^6, 7^ The two trials will enroll patients with severe degenerative MR, but have differences in age eligibility requirements (18+ versus 65+), surgical risk (moderate versus across the spectrum), and outcomes, as well as the time period over which these outcomes are evaluated. One trial will assess outcomes over two years, while the other trial focuses on longer-term outcomes over five to ten years.^6, 7^

A prior study examined patient preferences for a “beating heart” approach versus an “open heart” approach for MR repair; however, its sample size was limited (n <100); it did not include mortality risk as an attribute; and it focused on shorter-term risks (over 30 days and two years).^22^ That study found that patients were willing to accept an additional 10% in two-year risk of re-intervention for a less invasive approach (with less intensive recovery), holding all other factors equal. In comparison, our study focused on longer-term risks and benefits (i.e., over five years) and found that patients were willing to accept a slightly larger (13%) risk of re-intervention over a five-year time frame. While a transcatheter procedure, like TEER, is considered to be less invasive and have a shorter recovery time than surgery, understanding to what degree patients value these advantages compared to other factors, such as mortality risk, improvements in physical functioning, hospitalizations, and risk of needing re-intervention, will help ensure patient-centered treatment decisions.

Secondary analyses exploring heterogeneity in patient preferences identified three meaningful preference patterns, each representing approximately one-quarter to one-third of the respondent population. One group cared most about improving their physical functioning, the second group cared most about minimizing five-year mortality risk and secondarily re-intervention risk, and the third group cared most about receiving a transcatheter procedure over surgery. These findings are important preference phenotypes to keep in mind, as individual patients will likely prioritize different features to varying degrees. The identification of these preference phenotypes also suggests that in patients who are eligible for both interventions, a shared decision-making process that takes into account individual patient preferences will be important.

### Limitations

One limitation of all stated-preference research, including DCE surveys, is that they are based on self-report and rely on participants comprehending and being attentive to the questions. Another limitation is that participants were asked to consider a medical decision that they were not currently facing. We acknowledge that additional real-world factors may influence actual medical decisions. But, studies such as ours are valuable in providing a patient’s unadulterated views on acceptable benefit-risk tradeoffs using a controlled experiment. To ensure that the survey was clear, comprehension of the descriptions and graphics (e.g., risk grid tutorials) used in the survey were evaluated during pretesting, and the survey was improved in an iterative fashion. The survey also included comprehension questions and if participants answered these questions incorrectly, the correct answer would be presented along with an explanation. Thus, these comprehension questions served both as checks as well as training exercises for participants. Additionally, a series of internal validity tests were included in the study (e.g., scope test) and prior to analysis of response data, all of these internal validity tests were assessed (e.g., straight-lining). Another limitation of the study was the lack of diversity in recruitment, limiting the generalizability of study findings to those represented in the participant population.

This study found that on average, patients preferred a transcatheter procedure over surgery to the extent that some increase in risks or more hospitalizations were acceptable tradeoffs. To undergo a less invasive procedure for MR repair versus surgery, patients were also willing, on average, to sacrifice the degree to which their physical functioning would improve. If their current level of physical functioning was equivalent to NYHA class III, they would accept an improvement to a level of physical functioning similar to NYHA class II rather than an improvement to NYHA class I. On the other hand, if improvements in physical functioning with an intervention could improve physical functioning from NYHA class III to NYHA class I over five years, this gain in utility would offset negative preferences for surgery. If surgery also reduces the incidence of re-intervention and/or mortality versus a transcatheter approach, the level of improvement in physical functioning required to compensate for the more invasive procedure would shrink toward an improvement from NYHA class III to NYHA class II. Therefore, it is necessary to consider the combination of advantages and disadvantages of transcatheter versus surgical MR interventions.

When mature clinical trial data comparing outcomes with these interventions are available, findings from this preference study can be used to evaluate the average net benefit of alternative MR interventions. However, as our findings from latent-class analysis show, preferences for expected benefits and risks can vary significantly across individual patients. Preferences varied in regard to the relative importance of the type of intervention and patients’ prioritization of maximizing physical function benefits or reducing the risk of mortality risk or the need for re-intervention. An efficient, structured approach to eliciting preferences and individualized risk predictions could be valuable to facilitate discussion and to document understanding of benefit-risk tradeoffs in delivering patient-centered care.

## Data Availability

Data can be made available by contacting the study authors.

## Acknowledgments

We are grateful to the patient advisory group that was recruited through Heart Valve Voice who provided valuable feedback throughout the study.

## Author contributions

All authors meet the International Committee of Medical Journal Editors criteria for authorship. AH drafted the article. All authors made substantial contributions to conception and design of the manuscript and interpretation of data, revised critically for important intellectual content, and gave approval of the final version.

